# A Cross-Sectional Study Evaluating Tick-borne Encephalitis Vaccine Uptake and Timeliness Among Adults in Switzerland

**DOI:** 10.1101/2021.02.04.21251135

**Authors:** Kyra D. Zens, Vasiliki Baroutsou, Philipp Sinniger, Phung Lang

## Abstract

**Objectives:** The goal of this study was to evaluate timeliness of Tick-borne Encephalitis vaccination uptake among adults in Switzerland.

**Methods:** In this cross-sectional survey, we collected vaccination records from randomly selected adults 18-79 throughout Switzerland. Of 4,626 participants, data from individuals receiving at least 1 TBE vaccination (n=1875) were evaluated. We determined year and age of first vaccination and vaccine compliance, evaluating dose timeliness. Participants were considered “on time” if they received doses according to the recommended schedule ± a 15% tolerance period.

**Results:** 45% of participants received their first TBE vaccination between 2006 and 2009. 25% were first vaccinated aged 50+ (mean age 37). More than 95% of individuals receiving the first dose also received the second; ∼85% of those receiving the second dose received the third. For individuals completing the primary series, 30% received 3 doses of Encepur, 58% received 3 doses of FSME-Immun, and 12% received a combination. According to conventional schedules, 88% and 79% of individuals received their second and third doses “on time”, respectively. 20% of individuals receiving Encepur received their third dose “too early”. Of individuals completing primary vaccination, 19% were overdue for a booster. Among the 31% of subjects receiving a booster, mean time to first booster was 7.1 years.

**Conclusions:** We estimate that a quarter of adults in Switzerland were first vaccinated for TBE aged 50+. Approximately 80% of participants receiving at least one vaccine dose completed the primary series. We further estimate that 66% of individuals completing the primary series adhered to an “ideal” TBE vaccination schedule.

## Introduction

Tick Borne Encephalitis (TBE) is a severe central nervous system disease caused by the TBE virus and transmitted via infected ticks. TBE is among the most frequently diagnosed viral tick-borne diseases in Europe and both incidence and geographic range continue to increase[1]. While there are no curative therapies for TBE, two vaccines, Encepur and FSME-Immune, are available and recommended for individuals living, working or traveling within TBE-endemic areas[1].

As with many vaccines, TBE vaccination effectiveness is influenced by many factors. Age of first vaccination impacts initial immunogenicity and duration of protective responses. Adherence to priming and booster vaccination schedules is also essential. While less clear, TBE has two vaccine formulations and inconsistent use of a single vaccine type during priming may also affect immunogenicity. Although clear guidelines for TBE vaccination are in place, compliance by individuals/healthcare providers is not known. Such information is highly relevant for vaccination strategies and could improve effectiveness. The goal of this study was to evaluate adult TBE vaccination uptake in Switzerland, potentially identifying areas for improvement.

## Methods

To determine TBE vaccination coverage we conducted a national, cross-sectional study based on obtaining vaccination records by mail[2, 3]. Adults with a Swiss mailing address in each of three age groups (18-39, 40-59, 60-79) were selected from each of the 7 Swiss statistical “large regions”[4] by disproportional stratified random sampling. From each age group/region, 1,280 individuals were invited to participate (n=26,880). Individuals were requested twice by mail to submit a copy of their vaccination record. The study was approved by the Office of Data Protection and the Ethics Committee of the Canton of Zurich.

4,626 individuals submitted copies of their vaccination records (Table1). Records were manually inspected and number/date(s) of TBE immunization recorded. Data were adjusted for study design and non-response. For vaccine uptake analyses, participants with ≥1 TBE vaccination (n=1875) were evaluated. To estimate timeliness, individuals completing the primary series (≥3 vaccinations, n=1546) with complete vaccine type information were included (n=1178). Primary vaccinations were considered “on time” if they were received according to manufacturer’s schedules ± a 15% “tolerance period” (Table1). Booster vaccinations were considered “on time” if they were received within the 10-year interval recommended in Switzerland plus a 15% “tolerance period”. As schedules for Encepur and FSME-Immun differ, we selected the vaccine used to complete ≥2 of 3 priming doses for analysis. Analyses were performed using STATAIC16 and Prism 8. P values <0.05 were considered statistically significant.

**Table 1.**
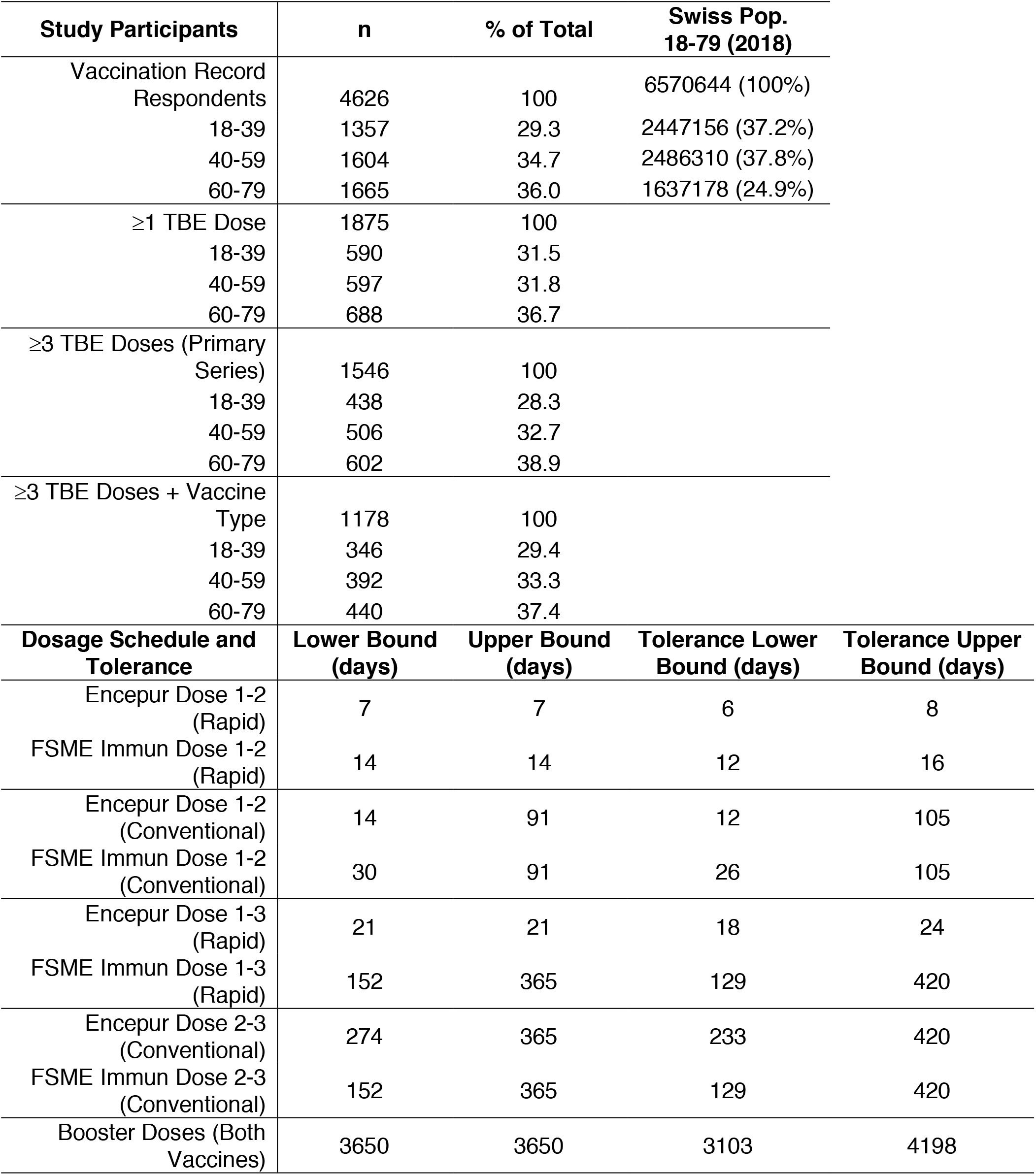
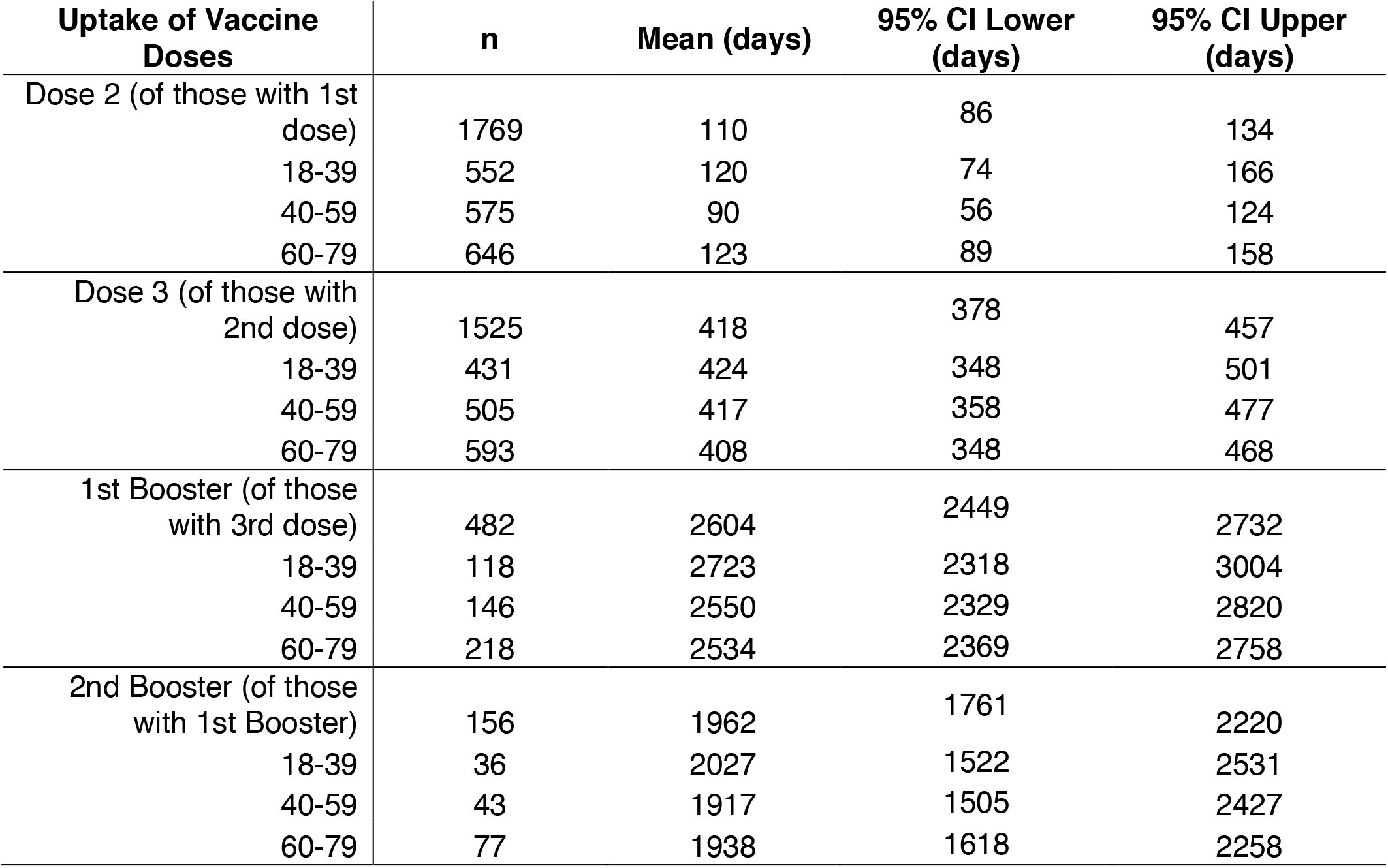
Study Participants, Dosage Schedule, and Days Between TBE Vaccine Doses.

## Results

Comparison of the year of first TBE vaccination among study participants showed a striking increase in uptake from 2006-2009, with 45% (95%CI 43-47%) of participants receiving their first dose (Figure1a). Mean age of first vaccination was 37. This varied significantly by age group; with a mean age of first vaccination of 22 for those 18-39, 40 for those 40-59 and 58 for those 60-79 (p<0.0001, Table1). 25% (CI 24-27%) of participants received their first vaccination aged 50+ (Figure 1b). Figure1c shows that 96% (95%CI 95-97%) of individuals receiving the first dose also received the second, with a mean time to vaccination of 110 days (95%CI 87-134; median 34 days 95%CI 33-35). 82% (95%CI 81-84%) of individuals receiving the second dose also received the third (mean time to vaccination 417 days, 95%CI 378-457; median 287 days 95%CI 282-294). We observed no difference between mean time to second or third doses between age groups (Table1).

**Figure 1.**
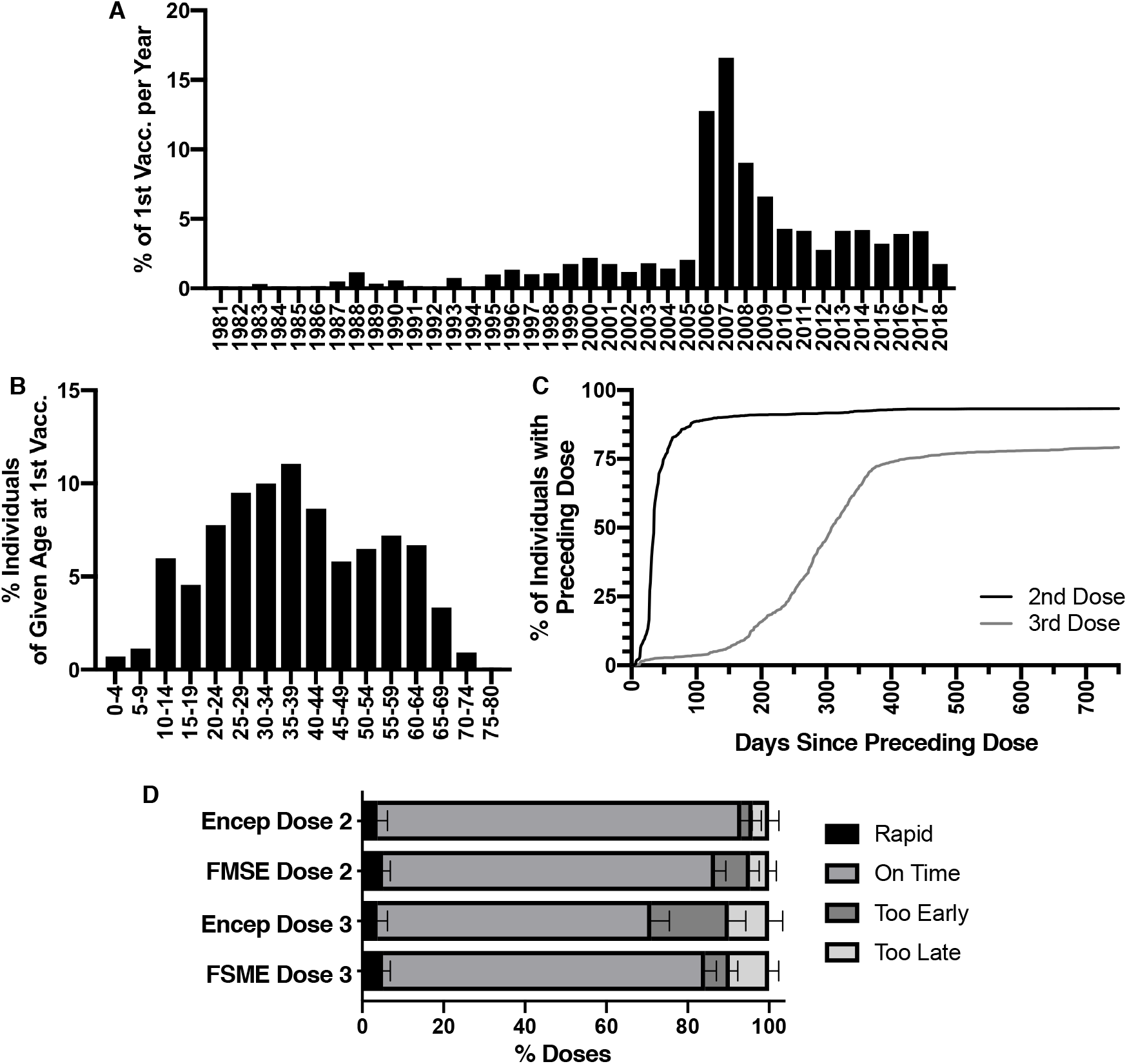
Year and Age of First TBE Vaccination. A) Year of First Vaccination: The frequency of individuals which received their first TBE vaccine dose in a given year, of individuals that received at least one TBE vaccine dose (n=1861, 14 values were excluded because of missing date information in the vaccination record). Note that 2018 was a partial year (study year). B) Age at First Vaccination: The frequency of individuals which received their first TBE vaccine dose at a given age, of individuals that received at least one TBE vaccine dose (n=1861, 14 values were excluded because of missing date information in the vaccination record, mean=37.2, 95% CI 36.7-37.6). C) TBE Vaccine Uptake: The percentage of individuals receiving the preceding dose which also receive the subsequent dose. (n=1854, 21 values were excluded because of missing date information for one or more doses in the vaccination record). D) Timeliness of Primary Vaccination by Vaccine Type: Among individuals completing the primary 3 dose TBE vaccination series with either majority Encepur (n=394) or FSME Immun (n=745), the percentage of 2^nd^ or 3^rd^ doses that were received as part of the Rapid Schedule, On Time, Too Early, or Too Late, based on the manufacturer’s recommendation plus or minus a 15% “tolerance period” (Table 1). Error bars represent one-sided 95% confidence intervals. Compared to the second dose, significantly fewer individuals received the third dose “on time” (78%, CI 76-81%, on time for dose 3 vs 88%, CI 86-91%, on time for dose 2, p<0.0001, Fisher’s exact test). 21% (CI 76-81%) of individuals receiving Encepur received their third dose “too early” compared to 6% (CI 4-8%) of those receiving FSME Immun (p<0.0001, Fisher’s exact test).

We further evaluated vaccine usage and adherence to recommended schedules. Among individuals completing the 3-dose primary series, 29% (95%CI 26-31%) received 3 doses of Encepur, and 59% (95%CI 56-62%) received 3 doses of FSME-Immun; 12% (95%CI 10-14%) received a combination. For both vaccines, 5% (95%CI 4-6%) of individuals followed “rapid” vaccination schedules (Figure1d). Of remaining individuals, 88% (95%CI 86-91%) received the second dose “on time”, based on “conventional” vaccination schedules, with no difference between vaccine types (Figure1d). Compared to the second dose, significantly fewer individuals received the third dose “on time” (78%, 95%CI 76-81%, Figure1d). Notably, 21% (95%CI 17-25%) of individuals receiving Encepur received their third dose “too early” compared to 6% (95%CI 4-8%) of FSME-Immun recipients (Figure1d). When we assessed adherence to a single priming vaccine type and “on time” completion of all priming doses together (including rapid schedule), 66% (95%CI 63-69%) of those completing the primary series adhered to an “ideal” schedule.

We then assessed uptake and timeliness of TBE booster vaccinations. Of individuals completing the primary series, 31% (95%CI 28-33%) received ≥1 booster(s). 19% (95%CI 17-22%) were overdue for a booster. Among those receiving a booster, the mean time between completion of the primary series and the first booster was 2590 days (95% CI 2449-2732, 7.1 years; median 2677 days 95%CI 2195-3142, 7.3 years). Second boosters were received significantly sooner; a mean of 1990 (95%CI 1761-2220, 5.5 years; median 1458 days 95%CI 1166-1840, 4.0 years; p=0.004) days after the first booster. There was no difference between age groups in mean time to first or second boosters (Table1).

## Discussion

Here, we found that 25% of adults received their first vaccination aged 50+. Age of first TBE vaccination impacts immune responsiveness and vaccine failures are increased among those 50+[5-7]. Interestingly, first vaccinations increased sharply in 2006, when Switzerland officially recommended TBE vaccination for individuals 6+ in many parts of the country[8], suggesting this prompted many individuals, including older adults, to be vaccinated.

Vaccination compliance can be evaluated by uptake and timeliness. While most participants receiving one TBE vaccine dose also received the second, uptake dropped between the second and third doses. Ultimately, ∼1 in 5 individuals beginning the primary series do not complete it. Regarding timeliness, nearly 90% and 80% of participants were “on time” for second and third doses, respectively. These findings suggest better compliance than German studies, where just over half of individuals initiating TBE vaccination completed the three-dose schedule[9] and less than one-third were vaccinated on schedule[10], or studies of other, non-TBE, adult vaccinees in the US and UK where compliance ranged between 30-50%[11, 12]. Whether this relatively high TBE vaccine compliance extends to other vaccines in Switzerland, though, is unclear.

Additionally, ∼20% of Encepur recipients received their third dose “too early”. This becomes only 5%, however, if the FSME-Immune schedule is applied (5-12 months rather than 9-12 months), suggesting that schedules for both vaccines are being used interchangeably. While it is unlikely “early” administration of Encepur negatively impacts immunogenicity, it indicates confusion regarding the vaccination schedule. As data support some interchangeability between vaccines[13], a unified recommendation for both vaccines may be warranted.

While manufacturers recommend TBE boosters every 3-5 years depending on age, TBE boosters are uniformly recommended every 10 years in Switzerland[8, 14]. In our study, mean time to first booster (7.1 years) was longer than manufacturer’s recommendations, but met Swiss recommendations[14]. In total, however, ∼1 in 5 participants completing the primary series was “overdue” for a booster suggesting the need for interventions to promote booster immunizations.

Here we demonstrate comparably high adult TBE vaccination compliance in Switzerland, though a substantial proportion of individuals were first vaccinated at an “advanced” age. A 2014 report estimated 4% vaccine failure among Swiss TBE cases[15]. We propose that studies of how irregular TBE vaccination and advanced age of first vaccination impact vaccine effectiveness are warranted to better inform vaccination policy decisions.

## Data Availability

Anonymized data used in this study can be made available upon request.

## Acknowledgements

We would like to thank Anna Fraefel, Carlotta Superti-Furga and Stefan Olarte for their help with data collection and organization during the study.

## Contribution

KZ analyzed data and wrote the paper, VB analyzed data, PS collected and analyzed data, PL provided financial support and supervision.

## References

1 Vaccines against tick-borne encephalitis: WHO position paper – Recommendations Vaccine. World Health Organization (WHO) 2011, pp 8769–8770.

2 Baroutsou, V., Zens, K. D., Sinniger, P., Fehr, J. and Lang, P., Analysis of Tickborne Encephalitis vaccination coverage and compliance in adults in Switzerland, 2018. Vaccine 2020. 38: 7825–7833.

3 Kantonales Durchimpfungsmonitoring Schweiz [Cantonal vaccination coverage surveillance Switzerland]. Swiss Federal Office of Public Health, Bern, Switzerland 2020.

4 Regional Statistics. Swiss Federal Statistical Office, Neuchatel, Switzerland 2020.

5 Weinberger, B., Keller, M., Fischer, K. H., Stiasny, K., Neuner, C., Heinz, F. X. and Grubeck-Loebenstein, B., Decreased antibody titers and booster responses in tick-borne encephalitis vaccinees aged 50-90 years. Vaccine 2010. 28: 3511–3515.

6 Andersson, C. R., Vene, S., Insulander, M., Lindquist, L., Lundkvist, A. and Gunther, G., Vaccine failures after active immunisation against tick-borne encephalitis. Vaccine 2010. 28: 2827–2831.

7 Hansson, K., Rosdahl, A., Insulander, M., Vene, S., Lindquist, L., Gredmark-Russ, S. and Askling, H. H., Tick-borne encephalitis (TBE) vaccine failures: A ten-year retrospective study supporting the rationale for adding an extra priming dose in individuals from the age of 50 years. Clin Infect Dis 2019.

8 Empfehlungen zur Impfung gegen Zeckenenzephalitis [Recommendation for Vaccination against Tick-borne Encephalitis], Bulletin 13 Edn. Swiss Federal Office of Public Health, Expert Vaccine Committee 2006.

9 Schley, K., Malerczyk, C., Beier, D., Schiffner-Rohe, J., von Eiff, C., Häckl, D. and Süß, J., Vaccination rate and adherence of tick-borne encephalitis vaccination in Germany. Vaccine 2021. 39: 830–838.

10 Jacob, L. and Kostev, K., Compliance with vaccination against tick-borne encephalitis virus in Germany. Clin Microbiol Infect 2017. 23: 460–463.

11 Johnson, K. D., Lu, X. and Zhang, D., Adherence to hepatitis A and hepatitis B multi-dose vaccination schedules among adults in the United Kingdom: a retrospective cohort study. BMC Public Health 2019. 19: 404.

12 Nelson, J. C., Bittner, R. C., Bounds, L., Zhao, S., Baggs, J., Donahue, J. G., Hambidge, S. J., Jacobsen, S. J., Klein, N. P., Naleway, A. L., Zangwill, K. M. and Jackson, L. A., Compliance with multiple-dose vaccine schedules among older children, adolescents, and adults: results from a vaccine safety datalink study. Am J Public Health 2009. 99 Suppl 2: S389–397.

13 Broker, M. and Schondorf, I., Are tick-borne encephalitis vaccines interchangeable? Expert Rev Vaccines 2006. 5: 461–466.

14 Schweizerischer Impfplan [Swiss Immunization Schedule]. Swiss Federal Office of Public Health, Bern, Switzerland 2020.

15 Schuler, M., Zimmermann, H., Altpeter, E. and Heininger, U., Epidemiology of tick-borne encephalitis in Switzerland, 2005 to 2011. Euro Surveill 2014. 19.

